# Magnitude of Needle Stick and Sharp Injuries and Associated Factors among Health Care Workers in Werabe Comprehensive Specialized Hospital, Southwest Ethiopia: A Cross-Sectional study

**DOI:** 10.1101/2022.11.16.22282377

**Authors:** Jemal Halil, Negalign Berhanu, Zeleke Dutamo

## Abstract

**Background:** Needle-stick and sharp injuries were one of major leading risk factor for blood and body fluid born infections among health care workers (HCWs)

**Objective:** To assess the magnitude of occupational needle-stick and sharp injuries and associated factors among HCWs in Werabe Comprehensive Specialized Hospital (WCSH), Southwest Ethiopia

**Methods:** Facility based cross-sectional study was conducted at WCSH from 15 to 25 August, 2020. Stratified random sampling was used to select 213 HCWs in the Hospital. A self-administered questionnaire was employed to collect data. After proportional allocation to the professionals, simple random sampling was used for each stratum. Data was entered and analyzed by using Epi info version 7 and SPSS version 22, respectively. Odds ratio was used to assess the statistical association between outcome and independent variables in bivariate and multivariate logistic regression. Significance of statistical association was tested using 95% confidence interval (CI) and P-value (<0.05)

**Result:** 28.40% of HCWs encountered needle-stick and sharp injuries in the last 1-year. HCWs who had job related stress, whose working hours was more than 8 hours per day and didn’t apply universal precaution were 8.6, 7.5 and 2.3 times more likely to encounter needle-stick and sharp injuries, respectively. HCWS with educational status above Diploma level were 90% less likely to face needle-stick and sharp injuries than their counterparts

**Conclusion and recommendation:** The preval**e**nce of occupational needle- stick and sharp injury was high compared to earlier studies. Educational status of diploma and above, average working hour for more than eight hours per day, not-applying universal precautions and job related stresses were factors associated with occupational needle-stick and sharp injures. Refreshment training on universal precaution and minimizing the excess working hours per day among HCWs were crucial to decrease the risk of needle stick and sharp injuries

## Introduction

Occupational needle stick and sharp injury refers to any injuries caused by medical set up sharp instruments that can cause scratch, cut or deep penetration to body parts during day to day activities in health care setting[1]. Needle stick and sharp injuries were one of the occupational hazards that can lead heath care workers (HCWs) to different infectious diseases, including more than 20 viral infections [2]. The commonest infections, that could be transmitted through needle-stick and sharp injuries are hepatitis B (HBV) & hepatitis C (HCV) and the human immunodeficiency virus (HIV)[3]

According to center for communicable disease control and prevention (CDC) estimate, 62% to 88 % of needle stick and sharps injuries in the hospital setting could be prevented by eliminating unnecessary injections, implementing universal precautions, eliminating needle recapping, provision and use of personal protective equipment, and training workers in the risk prevention [4, 5]

Each year, study reports in Africa, particularly Sub-Saharan including Ethiopia, indicated that large proportion of health care workers were exposed for occupational sharp injuries[6-9]

In Ethiopia, occupational needle-stick and sharp injury is still remain a challenge among HCWs, despite different efforts has been made by the government and non-government organizations on the implementation of safety. Very few studies were conducted to estimate the magnitude and associated factors of occupational needle stick and sharp injuries in Ethiopia [5, 9, 10]

So far, no study showed the magnitude and associated factors of occupational needle stick and sharp injuries in study area. Understanding of the magnitude and factors affecting the occurrence of occupational needle stick and sharp injuries enable NSI reduction; create safe working environment and provision of quality health care provision [11]. Therefore, this study tried to identify the magnitude and associated factors occupational needle stick and sharp injuries among health care workers in the study setting

## Methods

### Study design and setting

A facility based cross-sectional study design was employed. Study was conducted in Werabe comprehensive specialized Hospital. The hospital is found in Werabe town. It is an administrative capital of Siltie zone and located 172 km to the south of the country’s capital, Addis Ababa. The town has total population of 74,183. It has three urban and six semi urban administrative units (kebeles^a^). Werabe Comprehensive Specialized Hospital is a tertiary level hospital established in 2014, which was expected to serve 3.5-5 million populations and has a total of 800 beds. It provides a secondary and tertiary level of care and used as referral center for primary and general hospitals in the district and neighboring districts. A total of 700 employees were serving in different disciplines in the hospital. Out of 700 employees, 514 were HCWs including nurses (182), physicians (81), midwifes (56), laboratory technicians (45), cleaners (68), anesthetists (8), public health officers (10), psychiatry (9) and environmental health (6) professionals

### Sample size and Sampling procedure

Sample size was determined using single population formula considering 5% margin of error(d=0.05), 95%(z α/2_=_ 1.96) confidence level of certainty and 42% (p), proportion prevalence of needle stick and sharp injury conducted among HCWs in Arba Minch Referal Hospital, Southern Ethiopia and 10% of non-response rate by using formula

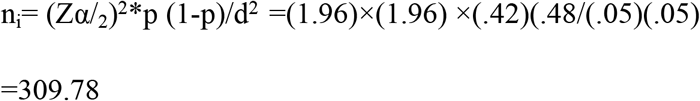

Since our source population 514 is smaller than 10,000, we used correction formula to obtain final sample size, i.e. n*f*= ni / (1+ni/N) =309.78/ (1+309.78/514) =193.29 ×10% (non-response rate) =212.62 ≈ 213

Hence, the sample size was 213

Stratified random sampling was used to select HCWs in the Hospital. HCWs were grouped into six main strata based on their occupation as nurses, midwifes, physicians, medical-laboratory technicians, cleaners and others (anesthetists, public health officers psychiatry and environmental health professionals). The required sample size for each stratum was determined by using proportionate allocation to the strata. Study participants from each professional category were selected by simple random sampling using lottery method (see Figure 1)

**Figure 1.**
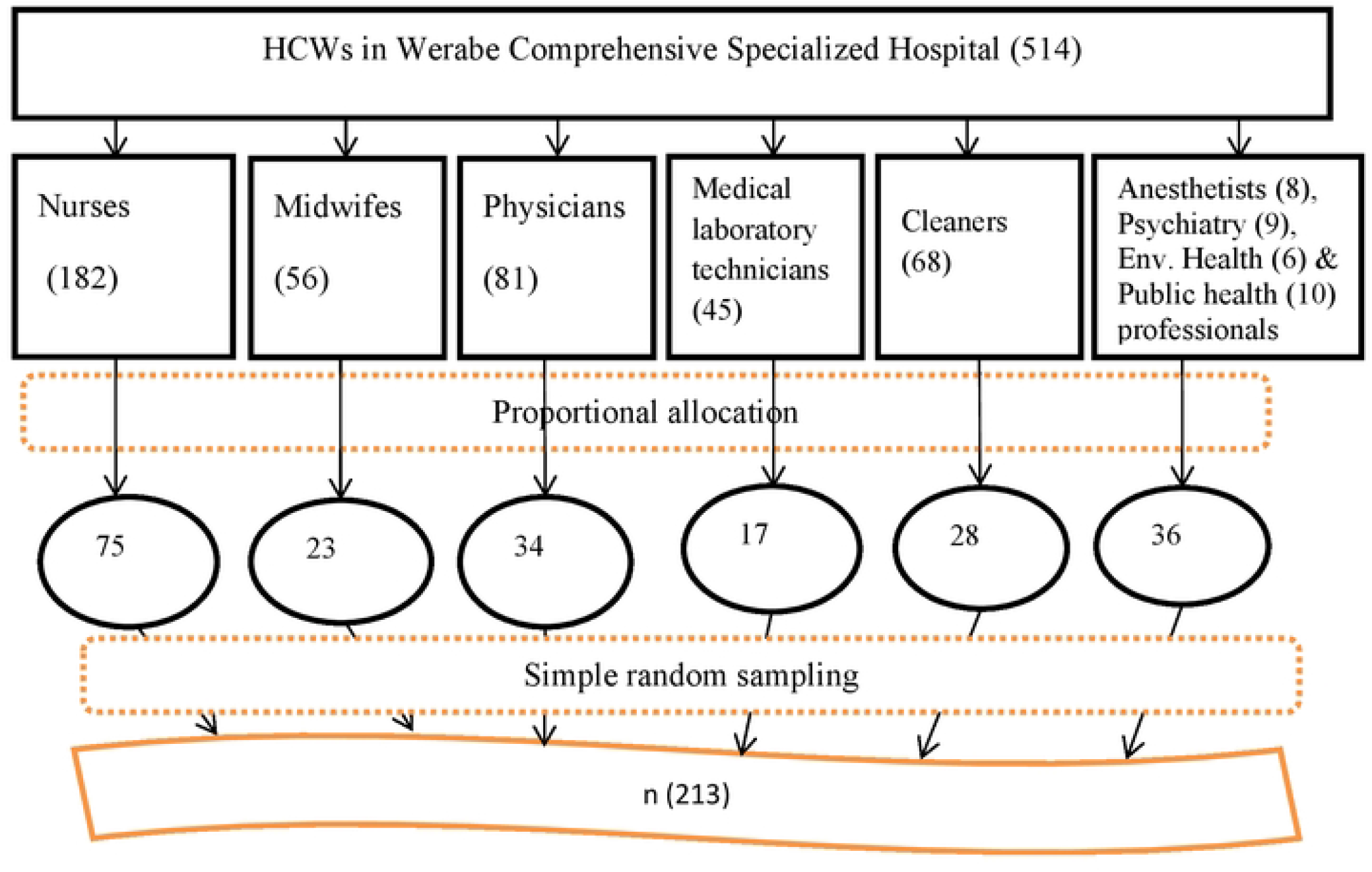
Schematic representation of sampling procedure to select HCWs in WCSH, Werabe, Southwest, Ethiopia, June, 2020

### Data collection tool and procedure

Data were collected by using structured self-administered questionnaire. The questionnaire was adapted by reviewing different literatures [5, 9]. Data were collected by three Diploma nurses. Questionnaire was first prepared in English and then translated into Amharic (working language) by a team containing language experts and health professionals. The Amharic version of the tool was retranslated into English in order to check consistency. In order to ensure data quality, two days training was provided to data collectors and supervisor. Pretest was conducted among 11 HCWs found in Kibet Primary Hospital. Close supportive supervision and assistance for data collectors was employed during data collection

### Ethical approval and consent to participate

Ethical clearance was obtained from Ethical Review Board of College of Medicine and Health Science, Wachemo University. An official letter of permission was obtained from Silte Zone Health department. Study participants were well informed on study purpose and the right to whether or not participate in the study. Full oral and written consent was obtained from all study participants. In order to maintain confidentiality of the study participants, anonymity of some personal identification were considered in the questionnaire

### Data processing and analysis

We entered data into EPi INFO version 7 statistical packages and analyzed by statistical Package for Social Sciences (SPSS) version 20. Study participants’ response to occupational needle-stick and sharp injuries status in the past 12months (outcome variable) was coded as “yes” and “No”. The Hosmer and Lemshow goodness of fit test was carried out and showed a p-value of 0.72 which indicated the model was adequate. Bivariate analysis was carried out using binary logistic regression analysis. Independent variables that had a p-value ≤ 0.25 in the Bi-variate analysis were included in the multivariable logistic regression model. A backward stepwise logistic regression analysis was carried out to control confounder for the predictor variable. The strength of association of predictor variables was assessed using adjusted odds ratio (AOR) and P-value <0.05 at 95% CI was considered statistically significant

## Result

### Socio demographic characteristics of the respondents

From a total of 213 HCWs, 204(95.8%) responded the questionnaire. One hundred nine (53.4%) of the study participants were females and the rest were males. One hundred thirty one(64.2%) of the study participants were in the age group of 25-29 years and the mean age of the study participants was 27.4(SD = ±3.9) years. Regarding to the marital status of the respondents, about half (51%) of the study participants were married. One hundred seventy one (83.3%) were Muslims by religion. One hundred twenty three (60.3%) of the study subjects were first degree and above in their educational status. The majority (68.0%) of the HCWs have less than five years of service. HCWs participated in the study had served in the hospital three and half years. Eighty one (39.9%) and forty three (21.1%) of the study participants were nurses and cleaners by profession, respectively (see Table1)

### Work environment related characteristics of study participants

From the total 204 respondents, one hundred thirty seven (67.2%) had not receive on the job training on infection perfection. The majority, 170 (83%) of the participants reported that there was a safety or universal precaution guidelines in their working place. One hundred thirty four (65.7%) of the respondents had received supportive supervision from concerned body on implementation of infection prevention and safety principles. One hundred seventy nine (87.7%) of the study participants reported that personal protective equipment were adequately provided by the hospital. More than half (55.4%) of the study participants experienced stressful situation related with their work or working environment. One hundred forty three (70.3%) of the study participants reported that, they were working for eight or less than eight hours per day and sixty one (29.7%) were working for more than eight hours per day (see Table 2)

### Prevalence of occupational needle-stick and sharp injuries

In this study, the prevalence of occupational needle-stick and sharp injuries was 28.4% (95% CI: 22.1-34.8). From the study participants who experienced needle-stick and sharp injuries, the majority (72%) did not report the incidence of the injuries to the concerned body

### Medical instruments causing needle-stick and sharp injuries

The main causes for needle-stick and sharp injuries were injection-needle (53.1%) and IV Cannula (17.2%). Moreover, 16% and 15% of the injuries were caused by surgical blade and suturing needles, respectively (see figure 2)

**Figure 2.**
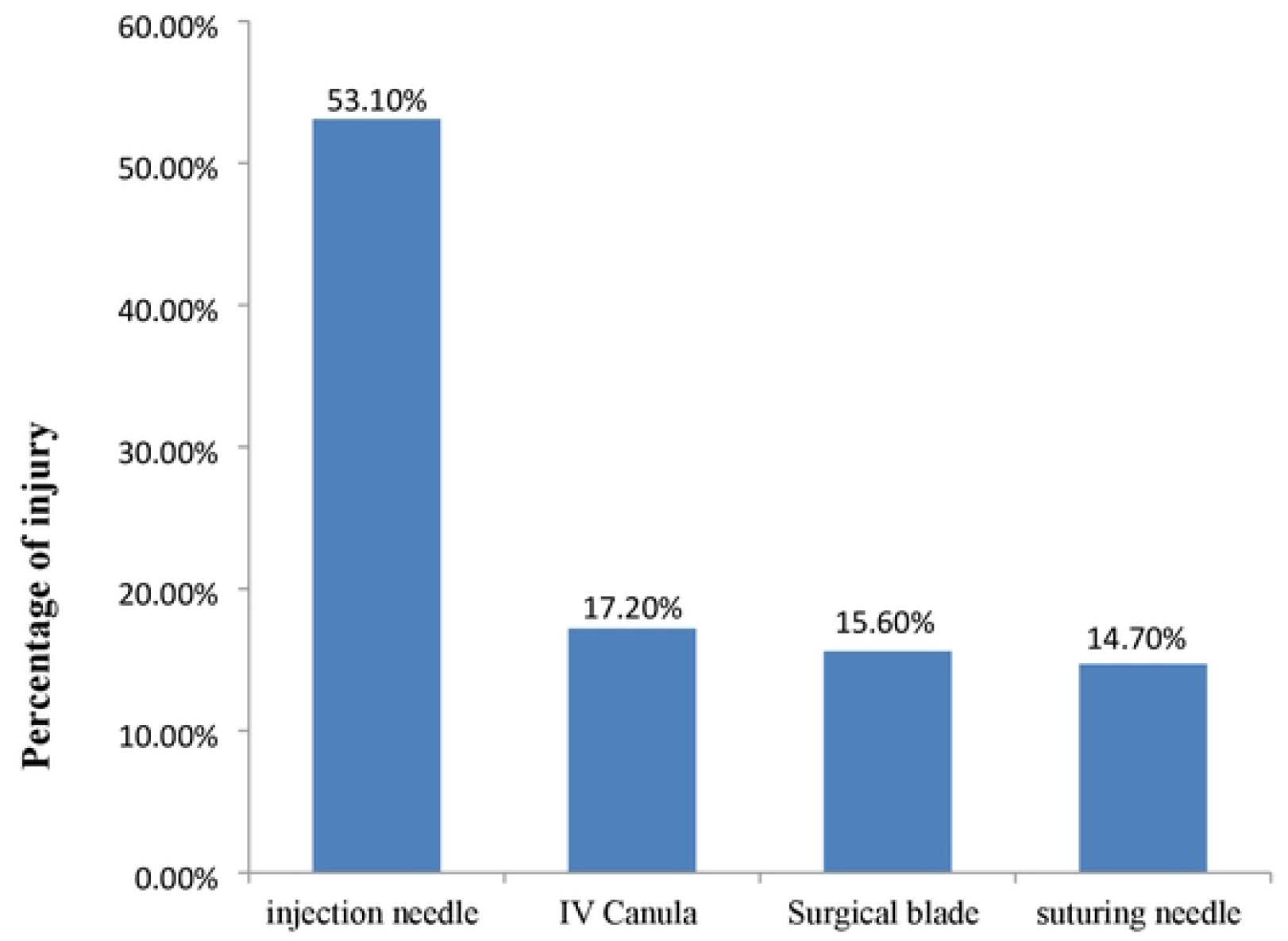
Medical instruments commonly causing needle -stick and sharp injuries among HCWs at WCSH, Werabe, Southwest Ethiopia, June, 2020 Figure 2 reasons why sharp injuries occurred among study participants at werabe comprehensive specialized hospital, June, 2020.

Among HCWs, about 41% and 30% of needle-stick and sharp injuries occurred during injection and sample collection procedure and medical waste handling, respectively (See figure 3)

**Figure 3.**
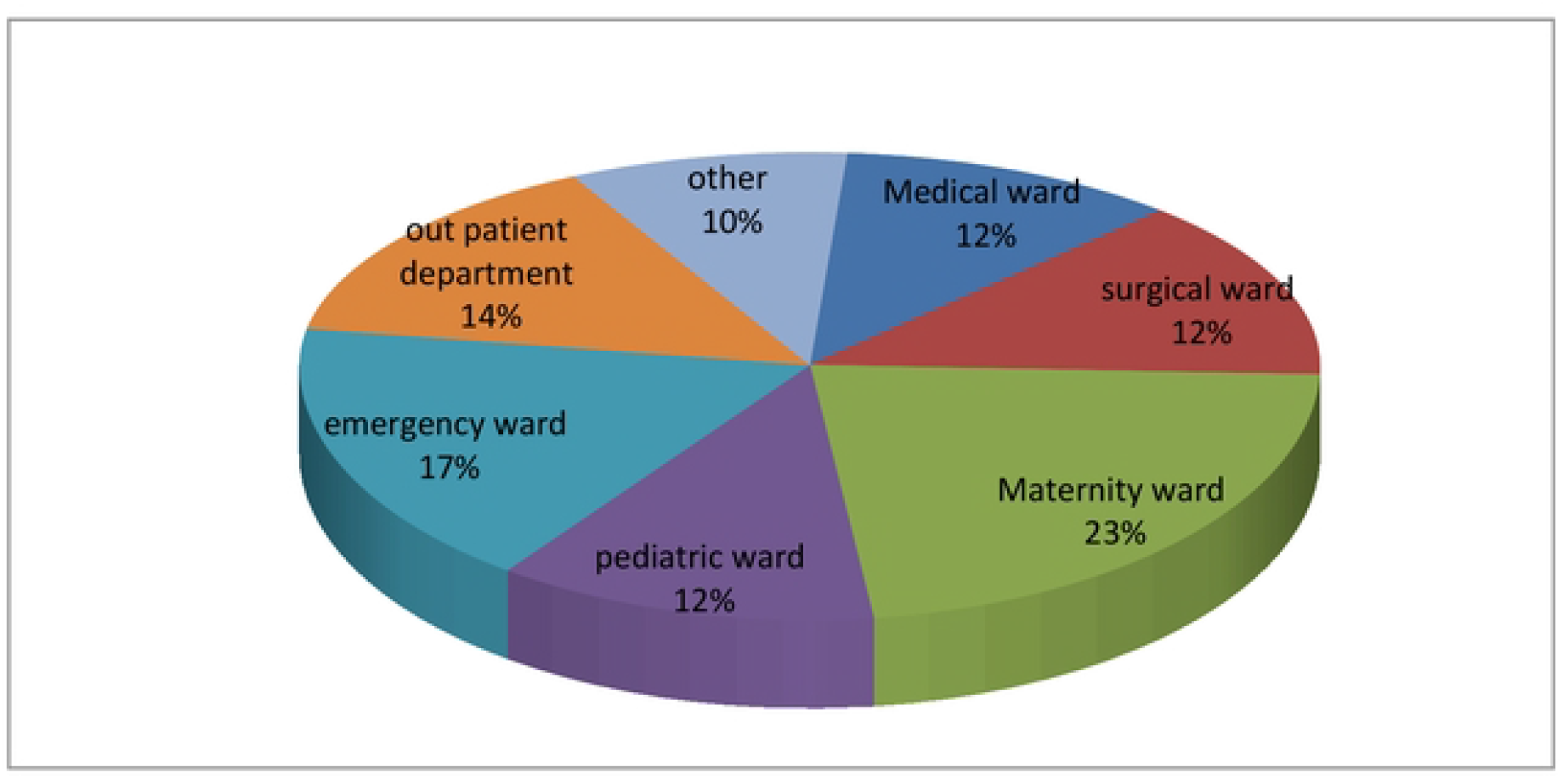
Percentage of occupational sharp injury among HCWs by their working department at werabe comprehensive specialized hospital, June, 2020.

**Figure 3.**
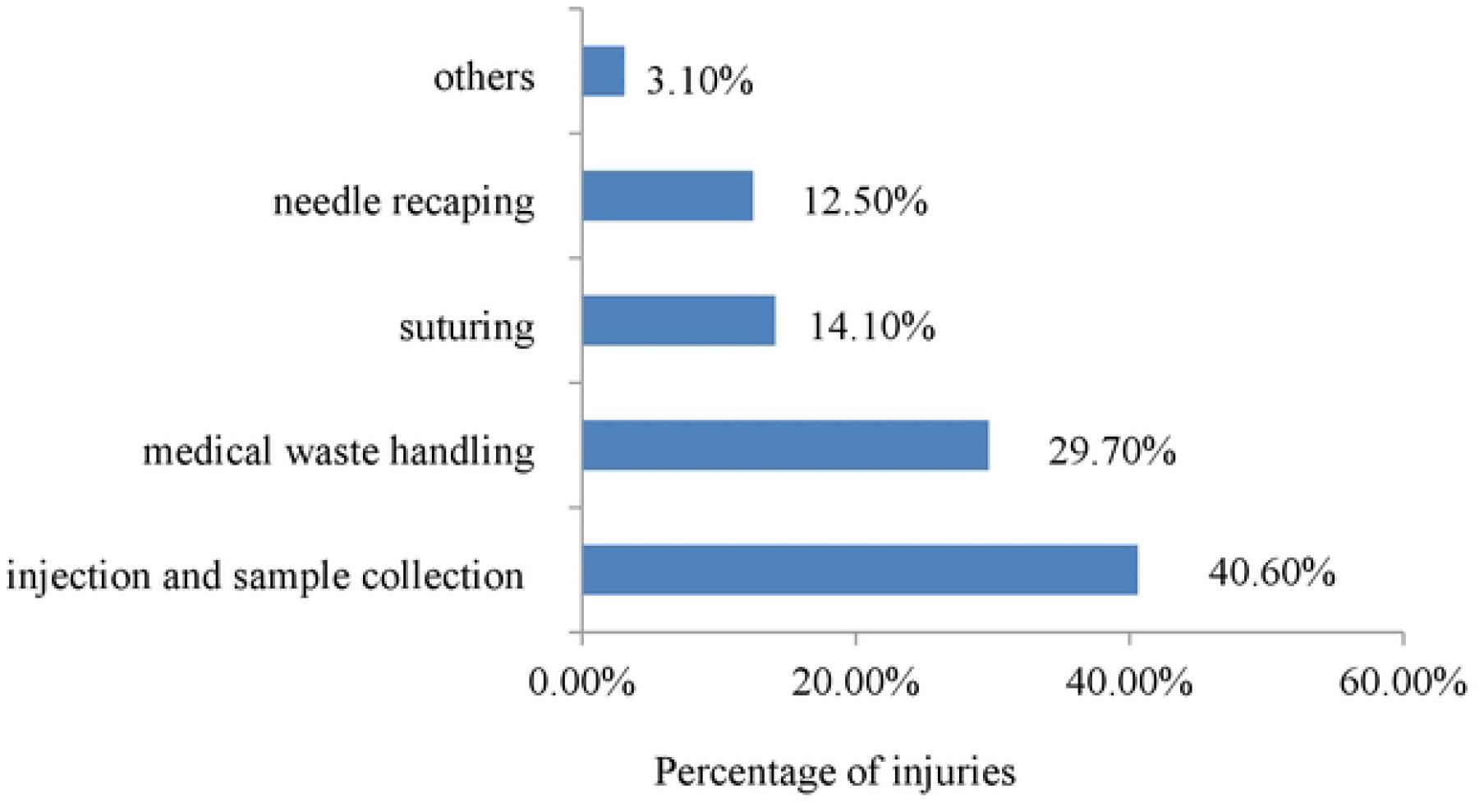
timeline when needle-stick and sharp injuries occurred among study participants at WCSH, Werabe, Southwest Ethiopia, June, 2020

Regarding with the working departments of the health care workers, 23%, 17% and 14% of all injuries occurred at maternity ward, emergency and outpatient department, respectively

### Factors associated with occupational needle-stick and sharp injuries

Educational status, average daily working hour, PPE use frequently and job related stress were significantly associated with occupational needle-stick and sharp injuries. HCWs with educational level of Diploma and above were 90% less likely to be injured than HCWs with educational level of 12^th^ grade and below (AOR 0.1, 95%CI 0.1, 0.5). Healthcare workers who worked for more than 8 hours per day had an almost 9-fold increase in risk of getting occupational needle-stick and sharp injuries than those who worked for eight and less hours in daily basis (AOR 7.5, 95% CI 3.8, 20.5). The study revealed that work related stress was one of the predictor of occurrence of occupational needle-stick and sharp injuries. The odd of injuries was about 5.6 times higher among stressed HCWs (AOR 5.6, 95% CI 2.4, 13.1) than who were not stressed. Inconsistent use of PPE increased the occurrence of occupational needle-stick and sharp injuries. HCWs who did not frequently use PPE in the last 12 months were about 2.4 times more likely to experience occupational needle-stick and sharp injuries (AOR 2.4, 95%CI 1.1, 5.4) than those who were frequently used PPE (see table 3)

## DISCUSSION

The result of this study revealed that the prevalence of occupational needle-stick and sharp injuries among HCWs in the last twelve months was 28.4%. The prevalence was higher when compared to study done in Saudi Arabia, Bale zone hospitals and Awi Zone, Ethiopia, where the prevalence was 6.4%, 19.4 and 18.7% respectively [10, 12, 13]. However, this finding was lower when compared to studies conducted in Tanzania, Nepal and Iran where the prevalence of occupational needle-stick and sharp injuries were 48.6%, 48.0% and 32.3% respectively [8, 14, 15]. The possible reasons for the difference could be access to use of advanced medical technologies and the length of the year at which studies were conducted. The type of HCWs participated in the studies may also increase or decrease needle-stick and sharp injuries prevalence

The current, study revealed that there was a significant association between educational level and occurrence of needle-stick and sharp injuries. HCWs with educational level of diploma and above were by 90% less likely experience needle-stick and sharp injuries than HCWs with educational level of grade 12^th^ and below. But, This finding differs from a previous study done in in Hawassa referral Hospital (Ethiopia), where HCWs with educational level of below diploma had less odds of injuries [16]. The possible justification might be as educational level of HCWs increased better practice of safety measures. The other reason could be less attention was given on work place supervision and on job training for those HCWs with lower educational level.

In this study statistical analysis showed that significant association between occurrence of sharp injuries and average daily working hour of the health care workers. HCWs who worked for more than eight hours were about 7.5 times exposed for occupational needle-stick and sharp injuries than those who worked for less than 8 hours on average in daily bases. This is in line with previous studies done in Iran, Addis Ababa (Ethiopia), Debrebrihan (Ethiopia) and Dessie (Ethiopia), where increased risk of sharp injuries occurred among HCWs who worked for more than 40 hours compared to HCWs worked less than 40 hours per week [17-20]. This implies that as the health care workers works for extended hour, they might experience a fatigue. This leads them less to adhere for safety protocols and might expose them for occupational sharp injuries.

In this study health care workers those who wet used personal protective equipment (PPE) like gloves during procedure were 2 times more likely at risk of getting sharp injury than their counterparts. This is in line with previous study done in Wolita zone public health facilities (Ethiopia) and Dessie town, northern Ethiopia [19, 20].inconsistent use of PPE during routine work by HCWs might be under estimation of the infection transmission by sharp instrument

Stressed HCWs were 5.6 times at risk of getting needle-stick and sharp injuries than who were not stressed. This is in line with previous studies done East Gojam zone, Ethiopia and Awi zone health institutions in north Ethiopia[12, 21]. This can be explained as HCWs who were stressed they become nervous, aggressive and might lose the ability of concentration during procedures. This might lead them to commit procedural errors. As a result they may expose themselves for preventable needle-stick and sharp injuries

## Conclusion

The result of this study showed that the prevalence of occupational needle-stick and sharp injuries was high. Educational status of diploma and above, working hour extended for more than 8 hours per day, not applying universal precautions and job related stress were predictors of occupational sharp injures. Refreshment training on universal precaution and minimizing the excess working hours per day among HCWs were crucial to decrease the risk of needle stick and sharp injuries

## Data Availability

Data will be accessed based on reasonable request from corresponding author

## End note

^a^ smallest administrative unit in Ethiopia

## Availability of data

Data set used and analyzed in this study available from the corresponding author and obtained based on reasonable request.

## Abbreviations and Acronyms

GP: General practitioner

HBV: Hepatitis B Virus

HCV: Hepatitis C Virus

HCW: Health care workers

HIV: Human immune deficiency virus

PPE: Personal protective equipment,

## Author contributions

JH took the lead role in conception, design, analysis and write up of the manuscript. ZD & NB involved in drafting the research proposal, analysis and write up of the manuscript. All authors read and approved the final manuscript.

## Acknowledgement

We would like to thank Silte District Health Department for financial support of the research undertaking. Also we would like to thank all respondents for their willingness to participate in the study.

